# Prevalence of Antimicrobial Residues in Tissues of Broilers Sold at a Local Market in Enugu State, Nigeria Using the European Four Plate Test

**DOI:** 10.1101/2021.09.19.21263803

**Authors:** O. S. Onwumere-Idolor, A. J. Ogugua, E. V. Ezenduka, J. A. Nwanta, A. Anaga

**Affiliations:** Department of Veterinary Public Health and Preventive Medicine, University of Nigeria, Nsukka, Nigeria; Department of Animal Health and Production Technology, Delta state Polytechnic, Ozoro, Delta State, Nigeria; Department of Veterinary Physiology and Pharmacology, University of Nigeria, Nsukka, Nigeria

**Keywords:** Antimicrobial Residues, Four Plate Test, Commercial, Broilers

## Abstract

**Background:** Consumption of animal tissues treated with antimicrobial agents may be deleterious if they contain violative levels of the residues. This study determined antimicrobial residues prevalence in broilers sold at Ikpa market, Nsukka, Nigeria.

**Methods:** Tissues from muscle, liver and kidney of 60 commercial broilers (180 samples) were tested for antimicrobial residues using the conventional Four Plate Test.

**Result:** Prevalence of 80.0% (48/60) and 51.7% (93/180) was recorded in the birds and samples {muscle 40.0%(24/60); liver 55.0%(33/60) and kidney 60.0%(36/60)} respectively. There was no significant association between residue occurrence and tissue type (χ^2^ (2) = 5.206; p = 0.074). Possible antimicrobial classes detected were: macrolid (50.0%) at pH 8.0 with *Micrococcus luteus;* β-lactams and tetracyclines (64.4%) with *Bacillus subtillis* at pH 6.0; sulphonamides (53.3%) at pH 7.2 with *Bacillus subtillis*, and aminoglycosides (46.8%) at pH 8.0 with *Bacillus subtillis*. Twenty-four tissue samples were positive at all four pH levels indicating use of more than one class of antimicrobials during each treatment.

**Conclusion:** Antimicrobial residues were detected in commercial broilers (muscles, liver and kidney) sampled at Ikpa Market, Nsukka. This is of public health importance given that antimicrobial residues are not monitored in poultry consumed in Nigeria.

## INTRODUCTION

The rise in demand for white meat has resulted in increased drive for poultry production in most parts of the world [1]. Poultry farmers therefore resort to the use of veterinary drugs, especially antimicrobials not only for therapeutic and prophylactic purposes but also as growth promoters to meet demand and make more money [2]. These antimicrobial drugs tend to accumulate in tissues as residues which could be consumed if the withdrawal periods are not observed. The presence of drug residues in meat above maximum residue limit (MRL), especially when withdrawal periods are not observed, has become a global issue. Lack of observation of withdrawal periods before processing animals for consumption is recognized worldwide by various public and government authorities as being illegal [3].This is because the consumption of meat with violative levels of antimicrobial residues could result in the development of antibiotic-resistant strains of microorganisms [4], allergic reaction in sensitised individuals, distortion of activities of the iintestinal flora, carcinogenesis and mutagenesis [5,6]. There is evidence of excessive prescription, overuse and abuse of antimicrobial drugs in veterinary practice in Nigeria, and legislations on the control of drug use in the country is hardly, if ever, enforced [2].

In Nigeria, antimicrobial residues in poultry meat are not regularly monitored as a matter of public health policy. It therefore becomes necessary to screen poultry meant for consumption in the country for the presence and levels of antimicrobial residues using reliable microbiological inhibition methods. These methods are less expensive when compared with immunochemical and chromatographic methods and yet are useful for detection of antibiotics or groups of antibiotics [7,8]. The Four Plate Test (FPT) is a microbial screening/inhibition test for detection of antimicrobial residues [9], in an intact tissue without clean up procedures and the determination of level of recoveries which are in immunological and chromatographic methods. The FPT is a four-plate agar diffusion test in which two different microorganisms; *Bacillus subtilis* and *Micrococcus luteus* are used as indicator organisms with three different pH levels of the media [10]. The aim of the study was to determine the prevalence and distribution of antimicrobial residues in broiler tissues sold at Ikpa market, Nsukka Enugu State, Nigeria using the European FPT.

## MATERIALS AND METHODS

### Study area and sample collection

Commercial broilers (60) were selected using simple random sampling from the list of broiler retailers in the Ikpa market, Nsukka. Only birds sold by retailers in the list were included. Birds sold by those not in the list were excluded. Birds sold in the market are sourced from poultries located within Nsukka and its environs. Two visits were made weekly and with six birds acquired per visit for 5 weeks (12 birds per week), a total of 60 commercial broilers were used. These birds were sacrificed humanely and from each, three tissue samples (muscle, liver and kidney) totaling 180 were harvested and subjected to laboratory analysis.

### Sample preparation

A 2g each of the harvested tissues was weighed, macerated and emulsified with equal volume of distilled water at a 1:1 ratio and introduced into centrifuge tubes. The tubes were centrifuged at 5000 rpm for 10 minutes and the supernatant decanted.

### Preparation of the FPT

Mueller-Hinton agar (MHA: Merck, Darmstadt, Germany) media was prepared in three different media bottles with pH adjustment from 7.4 to 6.0, 7.2 and 8.0 using HCL and NaOH to lower or increase the pH as required. The media was sterilized, allowed to cool, before dispensing into four corresponding petri dishes according to the pH. After gelling, the plates were seeded with *B. subtilus* (Merck Darmstadt, Germany, 64271) unto the first three plates with pH 6.0; 7.2 and 8.0. *M. luteus* (ATCC^®^ 10240) was seeded unto the last plate of pH 8.0, as instructed by Kilinc and Cakli [9]. These were stored for further analysis. Using a hole borer, four holes were bored on the surface of the agar plates and 100µl of the tissue extracts inoculated into the corresponding holes according to the tissue (muscle, liver and kidney). The fourth holes were inoculated with 100µl of distilled water and impregnated with an antibiotic as positive control. The plates were incubated for 24hours at 37°C and then viewed for the presence of inhibition zones (inhibition of the growth of the seeded test organisms) around the tissue samples and the control. A clear zone of inhibition with diameter ≥ 2mm was recorded as positive for antimicrobial residues.

### Statistical analysis

Data generated were analysed with Graphpad prism statistical package Version 5.2 for windows (Graphpad software, la Jolla California USA, www,graphpad.com) using Chi-square test to determine the association between categorical variables. Significance was accepted at α < 0.05.

## RESULTS

### Antimicrobial residues detected in the commercial birds

Out of the 60 birds screened for the presence of antimicrobial drugs residues, 48 (80%) were positive while the remaining 12 (20%) were negative as shown in figure 2.

**Figure 1:**
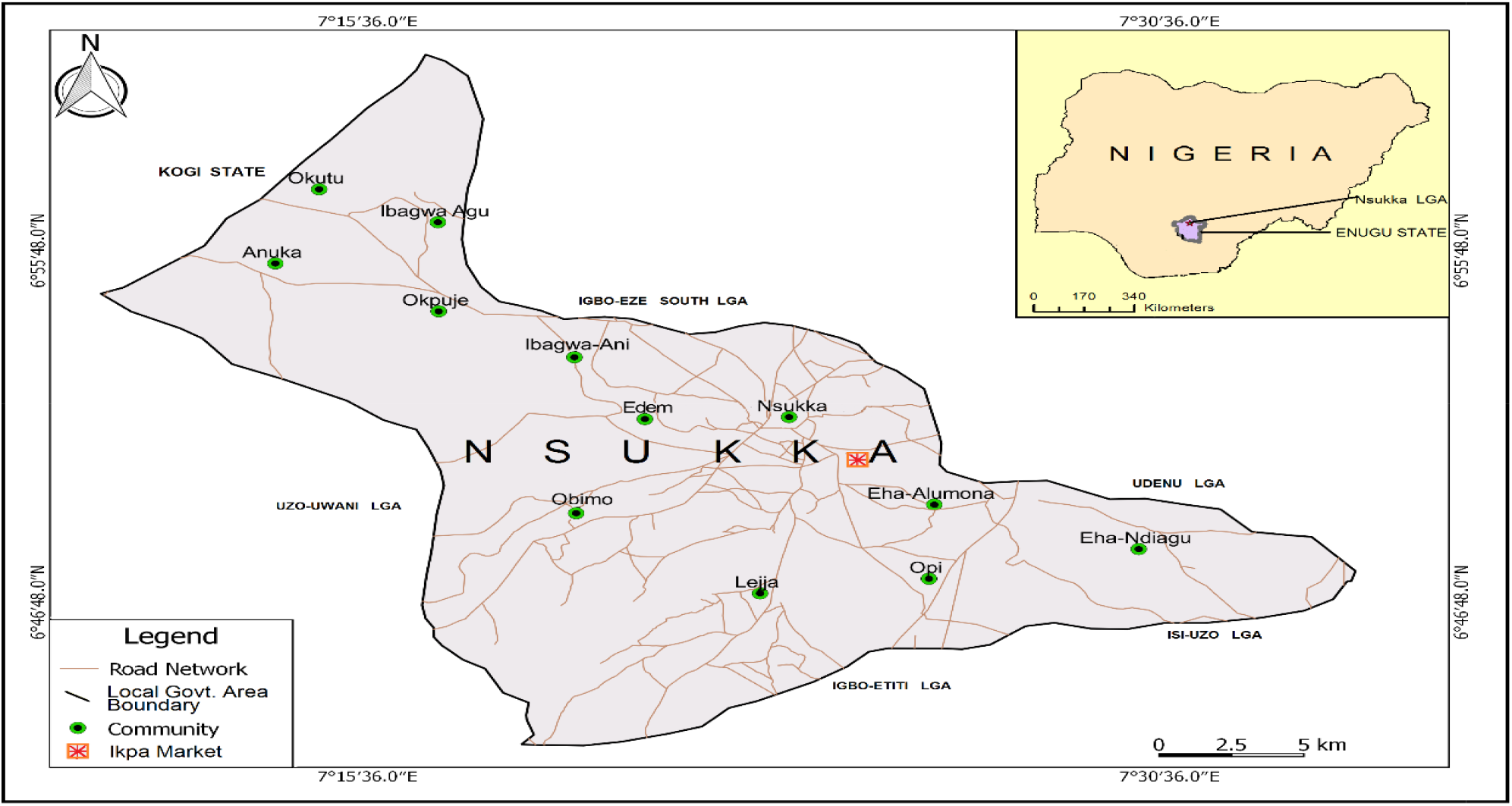
Map of Nsukka showing the point of sample collection

**Figure 2:**
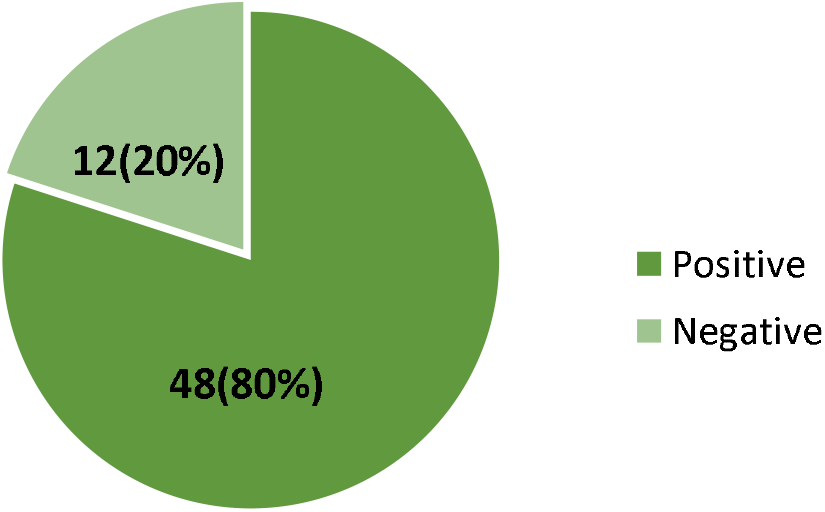
Prevalence of antimicrobial residues in 60 slaughtered commercial birds by FPT

### Antimicrobial Residues Detection in Sampled Tissues

Figure 3 shows the distribution of detected antimicrobial residues in the three tissues tested [muscle (24); liver (33) and kidney (36)]. There was no significant (χ^2^ (2) = 5.206; p = 0.074) association between antibiotic residues and tissue type.

**Figure 3:**
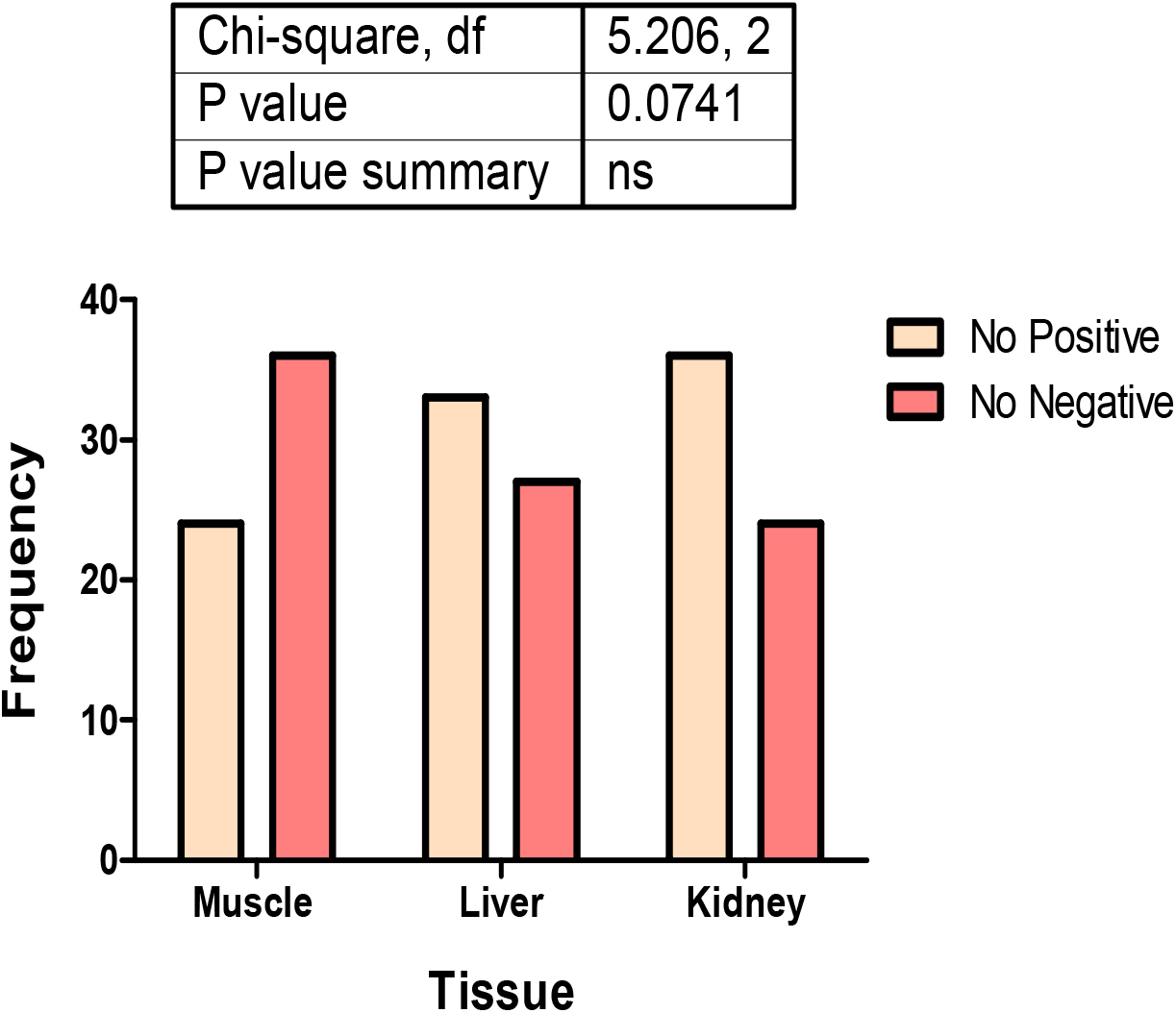
Tissue distribution of antimicrobial residues

### Possible classes of antimicrobials detected according to pH

The possible classes of antimicrobials detected from the different tissues tested were: macrolids (50%) at pH 8.0 with *Micrococcus luteus;* β-lactams and tetracyclines (64.4%) at pH 6.0; sulphonamides (53.3%) at pH 7.2 and aminoglycosides (46.8%) at pH 8.0 with *Bacillus subtillis*. The study detected more than one class of antimicrobial (multiple detection) in the tissues tested with the muscles having two (2), liver eighteen (18) and kidney four (4) (Table 1).

**Table 1:** Positive samples for different tissues at different pH levels.

| Tissues | Micrococcus | *Bacillus subtilis* (pH) |  |  |  |
| --- | --- | --- | --- | --- | --- |
|  | *luteus* pH 8.0<br>(%) | 6.0 | 7.2 | 8.0 | Multiple (all<br>four) |
| Muscle | 28 (46.7) | 30 (50) | 24 (40) | 12 (20) | 2 |
| Liver | 26 (43.3) | 46 (76.7) | 34 (56.7) | 30 (50) | 18 |
| Kidney | 36 (60) | 40 (66.7) | 38 (63.3) | 44 (73.3) | 4 |
| Total | 90 (50) | 116 (64.4) | 96 (53.3) | 86 (47.8) | 24 |

## DISCUSSION

The presence of drug residues in food is considered a public health hazard due to its deleterious effects to the consumers. This study found antimicrobial residues to be prevalent in the tissues of the birds screened. The prevalence of antimicrobial residues recorded in this study is higher than 60.0 and 33.1% recorded by Ezenduka *et al*. [3] and Kabir *et al*. [11] respectively, in different parts of Nigeria. Likewise, Ezenduka [12], recorded 64% prevalence while using Three Plate Test (TPT) which excluded pH 8.0 seeded with *micrococcus luteus* which has the best detectability for macrolids among plate tests. The differences in prevalence could be attributed to the fact that while this study used the FPT, the former two studies used the tube test and the latter one used the TPT. It is important to note that the conventional FPT has the capability to detect more antimicrobials including macrolids than the tube test. However, due to the fact that FPT is a qualitative microbiological test which basically is used for screening for the presence of antimicrobial residue [13], the test is not traditionally used to determine the exceedance of maximum residue limits (MRL). However, there are claims that screening tests (especially plate tests) detect the presence of antimicrobials when their concentrations are above MRL [14]. Similarly, prevalence of antimicrobial residues in broiler chicken has also been recorded in other developing countries 69.7% in Eastern province of Saudi Arabia by Al-Mustafa and Al-Ghamdi, [15], 70% in Pakistan by Jabbar [16], 52% in Iraq [17], 70% in Tanzania by Nonga *et al*., [18], 90% in Egypt by Karmi, [19] and 88% in Gaza Palestine by Abdelraouf *et al*. [20]. The occurrence of antimicrobial residues in foods of animal origin is indicative of excessive prescription, overuse and most importantly non-observance of withdrawal periods as asserted by the above-mentioned studies. Imprudent use of antimicrobials is common in Nigeria and has been attributed to the unrestricted availability of antimicrobial drugs and the practice of self-medication by most farmers in the country [21]. In the tissue distribution, the kidney and the liver had higher frequency of occurrence of antimicrobial residues than muscle tissue. This finding is in agreement with Pavlov *et al*. [22]. This may be as a result of the fact that while the liver is the organ of drug biotransformation and detoxification, kidney is the major excretory organ of most drugs.

Sixty-four percent of the antimicrobial residues were detected at pH 6.0 indicating possible β-lactams and tetracyclines. This is because pH 6.0 possibly detects two different classes of antimicrobials at the same time. The second highest detected was possibly sulphonamides (53.3%) class at pH 7.2, followed by the macrolids (50%) at pH 8.0 with *M. luteus*. The detection of the latter two classes may probably be as a result of the fact that the study was conducted during the rainy season when birds usually suffer from coccidiosis due to wet poultry environment making the oocyst to multiply. Birds suffering from coccidiosis are usually treated with sulphonamides based drugs while those having cough and respiratory diseases which are common in rainy season due to cold weather, are mostly treated with tylosin and other macrolids [23].

Detection of antimicrobials at different PH levels in the study suggests that different classes of antimicrobials were administered to the birds at the same time. Dipeolu [24] and Ezenduka *et al*. [3] found in their studies that broiler farms used different drugs at the same time to either treat or prevent diseases in their farms. The use of multiple classes of antimicrobials may be associated with the desperation of poultry farmers who use these drugs, to make up for poor biosecurity measures that expose their birds to infection, in the bid to curb the problem of concurrent infections in their farms [21]. This has very important implications for public health as this could give rise to the development of multidrug resistant microorganisms within the consumers. Infections due to multidrug resistant organisms are known to be difficult to treat, often requiring expensive antibiotics and long-term treatment that may be accompanied by side effects. This can substantially increase mortality and cost of treatment of infected persons/animals.

In spite of the results of the study, its limitation is that the sensitivity and specificity of the FPT microbiological method are generally low for residues quantification, but its simplicity and low cost makes it suitable for multiple antimicrobial residue screening test [25,26]. It is also more suitable for regulatory purposes and provides a greater accuracy in assessment of contamination level of poultry products with veterinary drug residues [11]. The test is therefore quite useful in developing countries.

## CONCLUSION

The study detected antimicrobial residues in commercial broilers screened. This is of public health importance given that the consumers of poultry meat in the study area are at risk of consuming violative level of antimicrobial residues. More drug residues were detected in liver and kidneys than tissues from other organs; therefore, care should be taken in the consumption of poultry offal in the study area. It is important to educate broiler farmers in the area on the necessity of observing withdrawal periods before selling their products for consumption. In addition, there is need for a national residue avoidance and control program in Nigeria.

## Data Availability

All data are included in the manuscript.

## Authors’ contributions

We declare that this work was done by the authors named in this article and all liabilities pertaining to claims relating to the content of this article will be borne by the authors. EVE, JAN and AA conceptualized the work; OSO and EVE conducted the experiments; acquired the data and drafted the manuscript; EVE analysed the data; AJO, OSO and EVE reviewed the manuscript and prepared it for publication; all the authors revised and approved the manuscript for publication.

## Acknowledgement

This study did not receive any specific grant from funding agencies in the public, commercial, or non-for-profit sectors.

